# Candidate Genes from an FDA-Approved Algorithm Fail to Predict Opioid Use Disorder Risk in Over 450,000 Veterans

**DOI:** 10.1101/2024.05.16.24307486

**Authors:** Christal N. Davis, Zeal Jinwala, Alexander S. Hatoum, Sylvanus Toikumo, Arpana Agrawal, Christopher T. Rentsch, Howard J. Edenberg, James W. Baurley, Emily E. Hartwell, Richard C. Crist, Joshua C. Gray, Amy C. Justice, Joel Gelernter, Rachel L. Kember, Henry R. Kranzler

## Abstract

**Importance:** Recently, the Food and Drug Administration gave pre-marketing approval to algorithm based on its purported ability to identify genetic risk for opioid use disorder. However, the clinical utility of the candidate genes comprising the algorithm has not been independently demonstrated.

**Objective:** To assess the utility of 15 variants in candidate genes from an algorithm intended to predict opioid use disorder risk.

**Design:** This case-control study examined the association of 15 candidate genetic variants with risk of opioid use disorder using available electronic health record data from December 20, 1992 to September 30, 2022.

**Setting:** Electronic health record data, including pharmacy records, from Million Veteran Program participants across the United States.

**Participants:** Participants were opioid-exposed individuals enrolled in the Million Veteran Program (n = 452,664). Opioid use disorder cases were identified using International Classification of Disease diagnostic codes, and controls were individuals with no opioid use disorder diagnosis.

**Exposures:** Number of risk alleles present across 15 candidate genetic variants.

**Main Outcome and Measures:** Predictive performance of 15 genetic variants for opioid use disorder risk assessed via logistic regression and machine learning models.

**Results:** Opioid exposed individuals (n=33,669 cases) were on average 61.15 (SD = 13.37) years old, 90.46% male, and had varied genetic similarity to global reference panels. Collectively, the 15 candidate genetic variants accounted for 0.4% of variation in opioid use disorder risk. The accuracy of the ensemble machine learning model using the 15 genes as predictors was 52.8% (95% CI = 52.1 - 53.6%) in an independent testing sample.

**Conclusions and Relevance:** Candidate genes that comprise the approved algorithm do not meet reasonable standards of efficacy in predicting opioid use disorder risk. Given the algorithm’s limited predictive accuracy, its use in clinical care would lead to high rates of false positive and negative findings. More clinically useful models are needed to identify individuals at risk of developing opioid use disorder.

**Key Points:** *Question:* How well do candidate genes from an algorithm designed to predict risk of opioid use disorder, which recently received pre-marketing approval by the Food and Drug Administration, perform in a large, independent sample?

*Findings:* In a case-control study of over 450,000 individuals, the 15 genetic variants from candidate genes collectively accounted for 0.4% of the variation in opioid use disorder risk. In this independent sample, the SNPs predicted risk at a level of accuracy near random chance (52.8%).

*Meaning:* Candidate genes from the approved genetic risk algorithm do not meet standards of reasonable clinical efficacy in assessing risk of opioid use disorder.

## Introduction

Opioid misuse and opioid use disorder (OUD) are significant public health problems. In 2022, 6.1 million Americans aged 12 or older met criteria for OUD,^1^ with 94.8% of these individuals acknowledging misuse of prescription analgesics and 40.9% reporting receipt of the medication from a healthcare provider.^1^ Given a surge in opioid overdose deaths,^2,3,4^ efforts have been made to identify individuals at greatest risk of opioid misuse.

However, at a population level, common genetic variation accounts for a small proportion of differences in OUD liability.^5^ Polygenic scores (PGS) derived from common genetic risk variants (i.e., single nucleotide polymorphisms, SNPs) across the genome account for much less variance in OUD risk (3.74%) than sociodemographic factors (41.32%).^6^ Nonetheless, attempts have been made to develop and commercialize genetic algorithms to identify individuals susceptible to developing OUD.^7-9^ These models usually include a few SNPs in genes that are considered causal candidates based on their presumed effect on neural systems that underlie OUD risk. In addition to their small effects,^10^ few candidate variants have been substantiated in genome-wide association studies (GWAS), a more rigorous method of identifying risk variants.^11-13^

Genetic predictive models are also vulnerable to confounding based on differences in patterns of genetic similarity, which can create the false appearance of strong prediction.^14^ If the prevalence of OUD varies among individuals with historically different geographic origins (either due to real differences or biases in the data used to train a machine learning (ML) model), the model will falsely attribute predictive power to SNPs that are markers of global or local similarity rather than disorder risk. Such spurious associations arise from population stratification, i.e., differences in allele frequencies that result from historical migration and mating patterns (Supplementary Figure 1). Used in real-world settings, these ML models are likely to bias predictions and lead to the false conclusion that they are useful for predicting risk of complex traits like OUD.

The U.S. Food and Drug Administration (FDA) recently gave pre-marketing approval to an algorithm that incorporates 15 SNPs with proprietary weights to predict OUD risk.^15^ We assessed these SNPs individually and collectively in relation to OUD in a genetically diverse sample of over 450,000 U.S. Veterans (Table 1) using logistic regression and ML analyses. Specifically, we assessed (1) whether the SNPs were individually associated with OUD risk, (2) how much variance in OUD risk the SNPs accounted for collectively, (3) whether the SNPs were better predictors of genetic similarity than OUD risk, and (4) whether basic demographics (i.e., age and sex) were better predictors of OUD risk than the SNPs.

**Table 1.** Opioid use disorder case/control status across genetically inferred ancestry groups.

|  | **Any Opioid Exposure** |  | **Short-term Opioid Exposure** |  |
| --- | --- | --- | --- | --- |
|  | **Cases** | **Controls** | **Cases** | **Controls** |
| ALL | 33669 | 418995 | 3704 | 121810 |
| EUR | 20392 | 284986 | 1982 | 82859 |
| AFR | 9496 | 85118 | 1211 | 22910 |
| AMR | 3215 | 39790 | 411 | 12586 |

*Note:* ALL = full sample, including individuals genetically similar to the East Asian and South Asian superpopulations, and those missing on genetically-inferred ancestry, EUR = individuals genetically similar to the European superpopulation, AFR = individuals genetically similar to the African superpopulation, and AMR = individuals genetically similar to the admixed American superpopulation.

## Methods

### Participants

The Million Veteran Program (MVP), an initiative of the U.S. Department of Veterans Affairs to discover the genetic etiology of diseases relevant to Veterans, was approved by the Central Veterans Affairs Institutional Review Board (IRB) and all site-specific IRBs. We followed all relevant ethical regulations for research with human subjects. Using electronic health record (EHR) data, we identified patients enrolled in MVP who had filled an outpatient prescription for any opioid analgesic. Cases were defined based on having at least one International Classification of Diseases (ICD)-9/10 OUD code. Controls were opioid-exposed individuals with neither an OUD diagnosis code nor any prescription fill for medications commonly used to treat OUD (i.e., buprenorphine, methadone, and buprenorphine/naloxone). This yielded 452,664 opioid-exposed individuals (M_age_ = 61.15, SD = 13.37; 90.46% male) with genotype data, including 33,669 cases. As sensitivity analyses, we removed cases who had received only one outpatient OUD diagnosis code (n = 8,355) and re-ran all models, which yielded similar results (Supplementary Tables). The sample’s genetically inferred ancestry (GIA) composition, which was assigned based on patterns of similarity to reference genomes of individuals in the 1000 Genomes project,^16^ was 67.46% European (EUR), 20.90% African (AFR), 9.50% admixed American (AMR), 0.81% East Asian (EAS), and 0.07% South Asian (SAS), with 1.25% unassigned (Supplementary Figure 2).

Among opioid-exposed individuals, we also selected a subset with only short-term documented exposure to opioids (4-30 days total), as this reflects the relative opioid naivete of patients for whom genetic testing would likely be sought. This yielded 125,514 individuals (3,704 cases) whose mean age was 59.98 (SD = 14.84) and who were predominantly male (90.37%). This subsample’s GIA composition was similar to the full sample (67.59% EUR, 19.22% AFR, 10.36% AMR, 1.25% EAS, 0.11% SAS, and 1.47% unassigned).

### Genotyping and Imputation

MVP samples were genotyped using a custom Affymetrix Axiom Biobank Array (MVP Release 4). Quality control and imputation were performed by the MVP Genomics Working Group.^17^ Duplicate samples and those with a sex mismatch, seven or more relatives in MVP (kinship > 0.08), excessive heterozygosity, or a genotype call rate < 98.5% were removed. Monomorphic variants and variants with high missingness (call rate < 0.8) or a Hardy– Weinberg equilibrium *p-*value < 1×10^−6^ were removed. Genotypes were phased with SHAPEIT4^18^ and imputed using Minimac4 software,^19^ with biallelic SNPs imputed using the African Genome Resources reference panel by the Sanger Institute.

To infer similarity to reference genomes and generate principal components (PCs) to account for global and local differences in genetic similarity, GIA composition was calculated by the MVP gwPheWAS Working Group.^20^ Reference genomes used to infer genetic ancestry were from individuals from the Yoruba in Ibadan, Nigeria (YRI), the Luhya in Webuye, Kenya (LWK), the British in England and Scotland (GBR), the Han Chinese in Beijing, China (CHB), and Peruvians from Lima, Peru (PEL). Further details on how subgroups were derived from genetic similarity PCA coordinates by the MVP gwPheWAS Working Group can be found in Hunter-Zinck et al.^17^ In summary, a random forest classifier was trained on the reference dataset using the first ten PCs. The algorithm was applied to the MVP principal components analysis (PCA) data, and GIA was inferred when the classifier’s predicted probability was greater than 50%. Individuals with lower assignment probabilities were retained in the full sample analyses. Although guidelines on the use of population descriptors in human genetics research^21^ were published after GIA analyses were conducted in MVP, we follow the recommendations as closely as possible.

### Single-SNP Association Analyses

We conducted association analyses for each of the 15 candidate SNPs with OUD case/control status using logistic regressions in PLINK 2.0.^22^ Analyses were also performed within GIA groups and the subset of short-term opioid-exposed individuals. To account for non-independence, we randomly removed one individual from each pair of related individuals (N = 24,585). Three sets of analyses were conducted: (1) with no covariates, (2) including the first 10 genetic similarity PCs, and (3) including age, sex, and genetic similarity PCs as covariates.

### Combined-SNP Regression Analyses

Using the glm() function in R, we fit logistic regressions to examine the collective association of the 15 SNPs with OUD case/control status. We followed a procedure similar to that used with the single-SNP models, performing analyses in the full sample, short-term opioid-exposed subsample, and GIA groups. We also ran three sets of models with increasing levels of adjustment, akin to the single-SNP models. We calculated Nagelkerke’s R^2^ to estimate the proportion of variance in OUD status accounted for by each model and the area under the receiver operating characteristic curve (AUC-ROC) to evaluate performance.

### Machine Learning Models

We developed ensemble ML models in the full sample, the subset of short-term opioid-exposed individuals, and within GIA groups. We chose to use ML techniques, as they are useful for modeling non-linear relationships between variables. However, their performance is constrained by the strength of the underlying genetic effects on OUD risk.

To examine ML’s ability to predict OUD vs. genetic similarity, additional models were developed using data confounded by inferred genetic ancestry, such that all cases were in one GIA group and controls in another.^14^ The confounded models show how biases in the patterns of population stratification within training datasets can influence model performance. All ML analyses were performed using the *caret* package in R, which allows for consistency in the data structure and preprocessing steps across models.^23^

To prepare the analysis, dosage data for each of the 15 SNPs were recoded as hard calls (i.e., “0”, “1”, or “2”), reflecting the number of risk alleles for each individual. Of the 15 genetic variants of interest, 8 were directly genotyped and 7 were imputed, all with imputation quality > 0.8. In all models, controls were under-sampled to yield equal proportions of cases and controls, which was required to address the severe imbalance in the number of cases relative to controls. In confounded models, we sought to preserve the greatest number of cases. Thus, in all models where GIA was unbalanced, the more common GIA group was assigned as cases and the less common one as controls. We used a 75%/25% data split to obtain independent training and testing sets. To prevent overfitting, we used 10-folds cross-validation during training, which systematically validates the model across subsets ("folds”) of the data. We used *caret*’s default tuning parameter search feature, with the optimal parameters selected based on ROC to provide the best balance between sensitivity and specificity.

We selected as base ML models two random forest implementations, “rf” (Breiman and Cutler’s random forests) and “ranger” (recursive portioning with random forests). We also used a linear support vector machine (SVM) implementation (svmlinear2, using the e1071 library^24^ in caret) to approximate the methodology used for the algorithm that received pre-marketing approval, as the actual algorithm is proprietary.^9^ Base model predicted probabilities were aggregated with the 15 SNPs to serve as predictors in a stacked ensemble ML model, which was trained using a binomial generalized linear model. This step allows the ensemble model to learn from the predictions of each of the base ML models, which can be used to enhance its performance. Accuracy, sensitivity, and specificity were the primary metrics used to evaluate model performance, although we also examined positive and negative predictive values as secondary metrics. These metrics were used to calculate a diagnostic odds ratio (DOR), indicating how much higher the odds are that the classifier produces a positive prediction in an individual with OUD than an individual without OUD.

## Results

In single-SNP models that did not account for genetic similarity, 13 of 15 SNPs were significantly associated with OUD risk after Bonferroni correction (Table 2). However, upon inclusion of measures of global genetic similarity (i.e., PCs) that number declined to three (Table 2). Five of the ten SNPs whose associations became nonsignificant when we controlled for genetic similarity had opposite directions of effect in analyses that were uncontrolled. In analyses within GIA groups that accounted for local variations in genetic similarity by including 10 PCs, the three significant SNPs were associated with OUD risk only in individuals genetically similar to the EUR superpopulation. Similar results were obtained among short-term opioid-exposed individuals (Supplementary Tables 1 and 2).

**Table 2.**
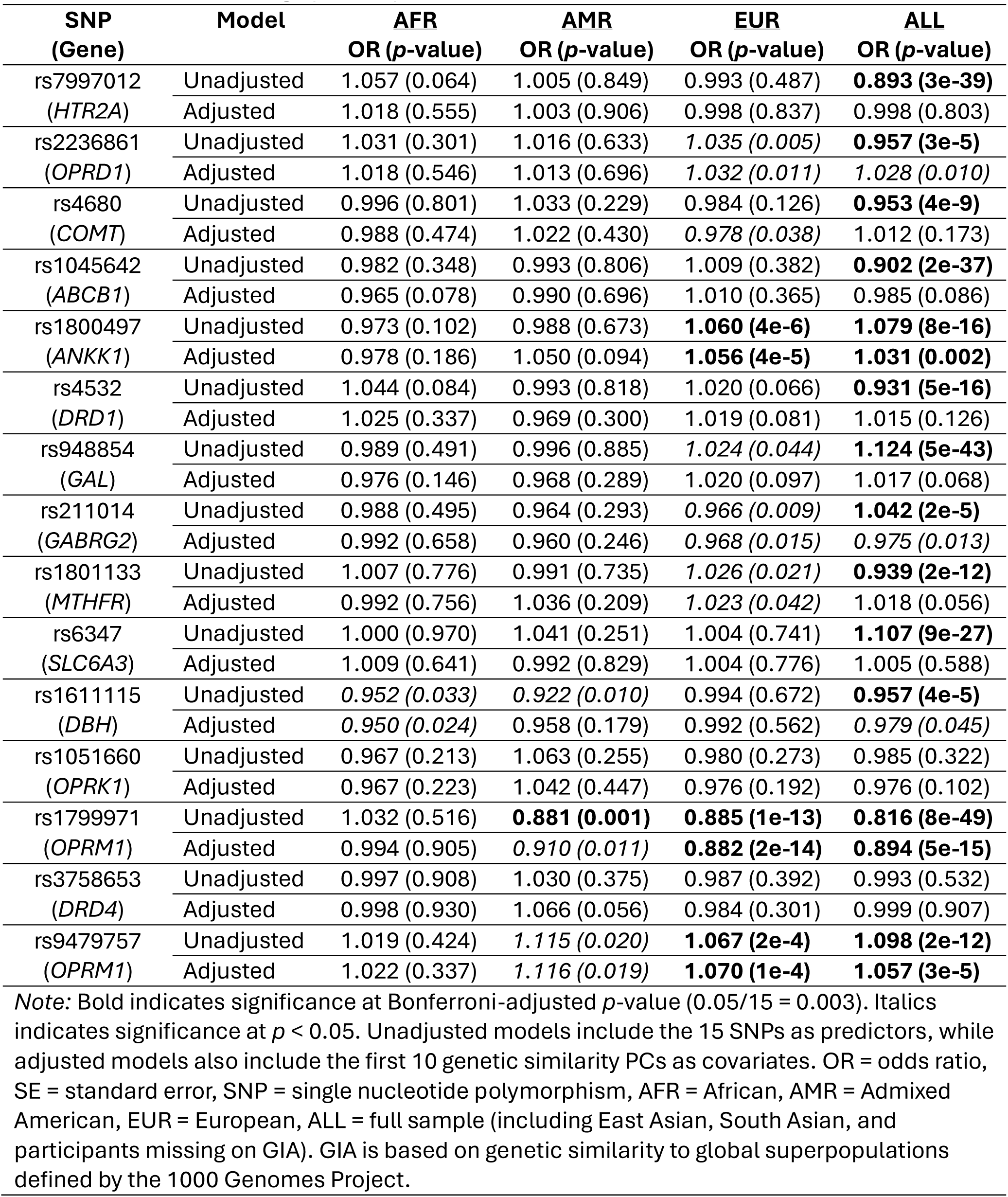
Single nucleotide polymorphism (SNP) associations with opioid use disorder case/control status among opioid exposed individuals.

In full sample logistic regressions, the 15 SNPs collectively accounted for 0.40% of the variance in OUD status (Figure 1), with an AUC of 0.54. In comparison, age and sex alone accounted for 3.27% of the variance and improved the model AUC to 0.66. Including PCs as covariates reduced the number of significant SNP associations from 11 to 5. In analyses conducted within GIA groups, 7 SNPs were significant in the EUR group, 2 in the AFR group, and 1 in the AMR group. The variance collectively accounted for by the SNPs was between 0.04% (AFR) and 0.16% (AMR) in models conducted within GIA groups, and AUC values ranged from 0.51 (AFR) to 0.53 (AMR). Supplementary Tables 3 – 6 provide full results of the logistic regressions.

**Figure 1.**
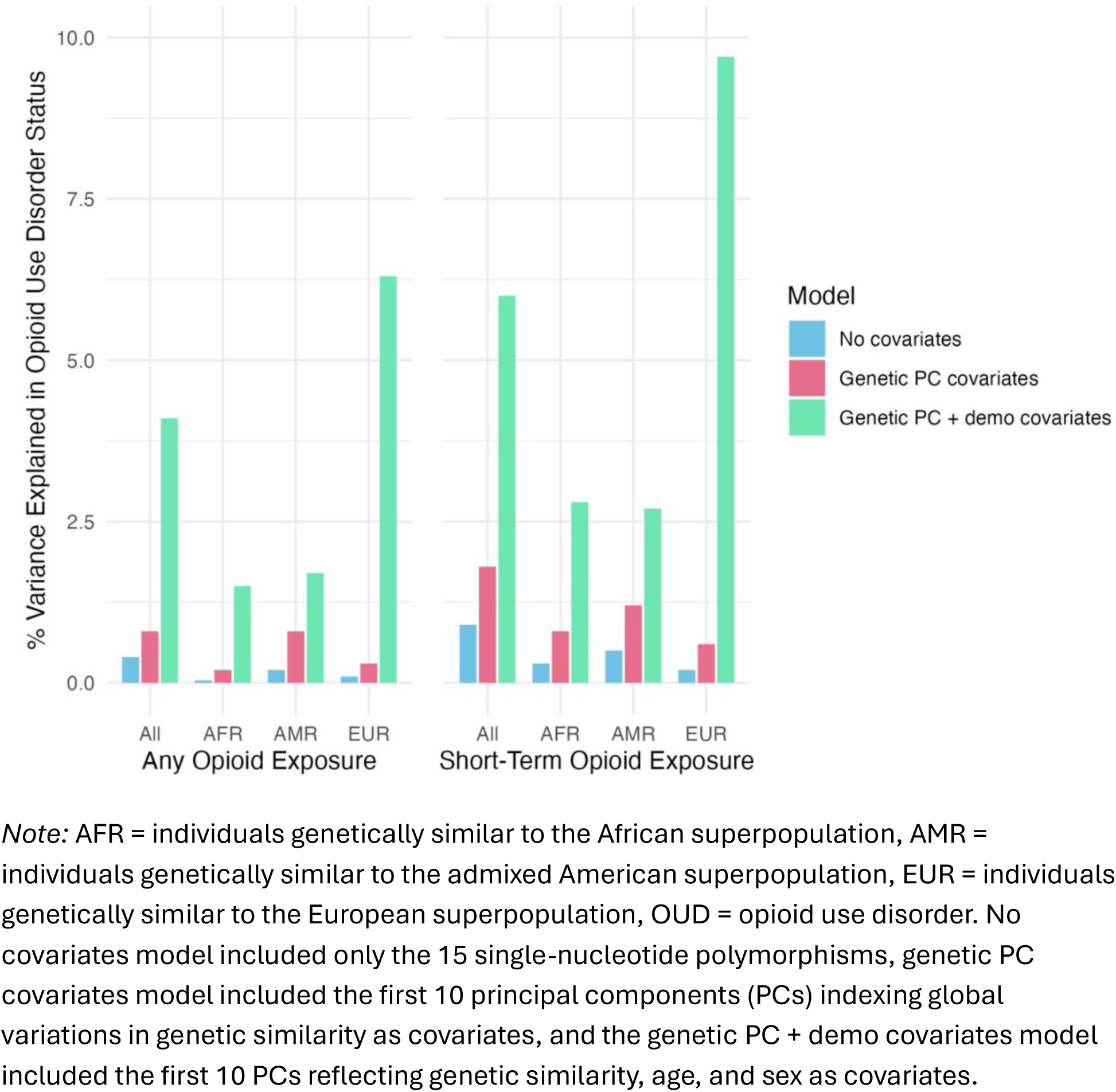
Percent of variance in opioid use disorder case/control status explained by combined-SNP regression models.

The accuracy of the ensemble ML model predicting OUD risk in the full sample (Figure 2) was slightly greater than chance (52.83%, 95% CI = 52.07 to 53.59%). In test data, the model correctly identified 50.72% of OUD cases (sensitivity) and 54.95% of controls (specificity). Of the model’s predicted cases, 52.96% were true cases (positive predictive value), and of the predicted controls, 52.72% were true controls (negative predictive value). The DOR was 1.25, suggesting that a predicted case is 1.25 times as likely to have OUD than a predicted control. In analyses conducted within GIA groups that accounted for local variations in genetic similarity by including 10 PCs, accuracy did not exceed random chance (EUR: 50.65% [95% CI = 49.67 to 51.62%]; AFR: 50.53% [95% CI = 49.09 to 51.96%]; AMR: 49.69% [95% CI = 47.21 to 52.16%]). Similarly, the DOR dropped to between 0.99 (AMR) and 1.03 (EUR). Similar results were obtained for the short-term opioid-exposed subsample (Supplementary Tables 7 and 8).

**Figure 2.**
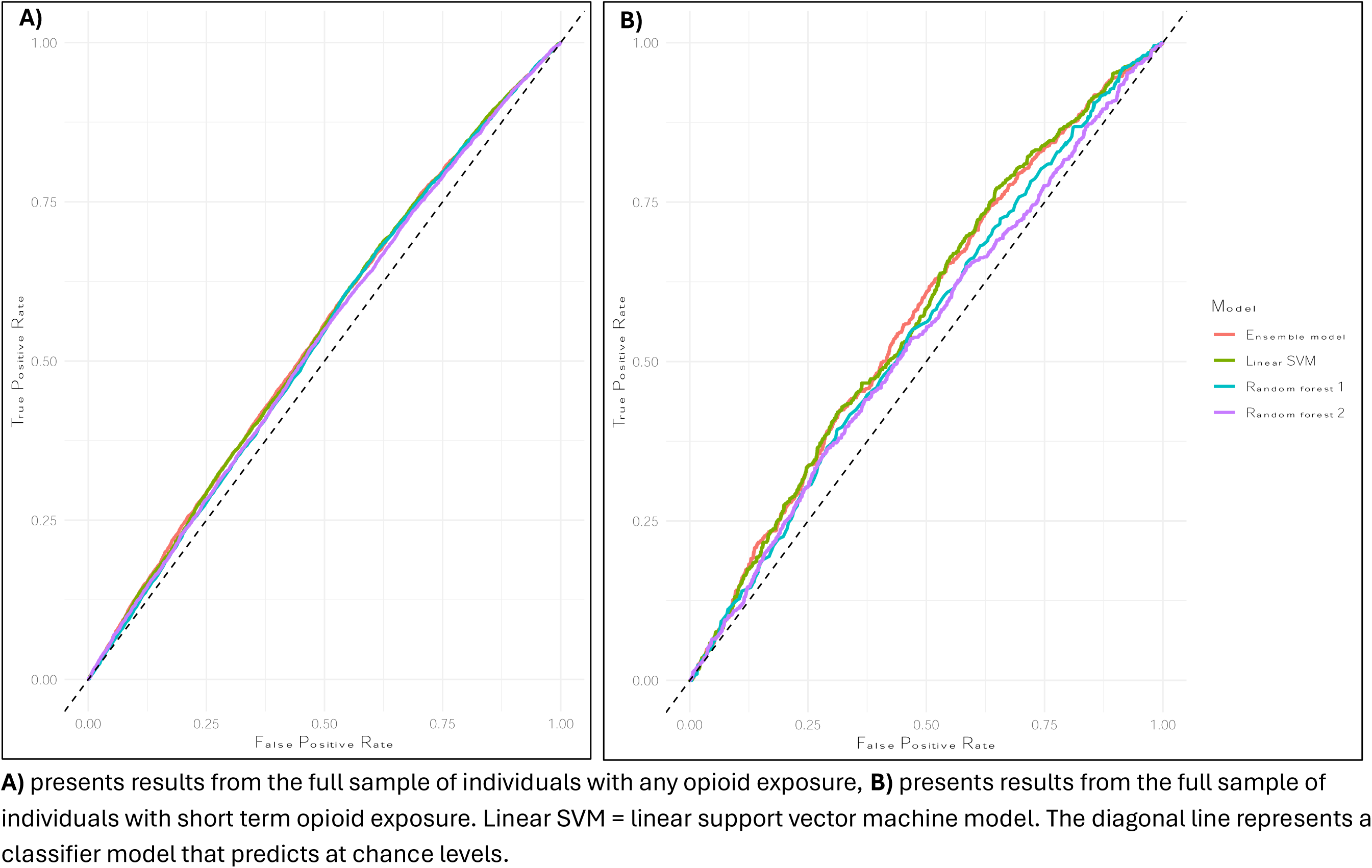
Receiver operating characteristic curves predicting opioid use disorder case/control status.

Confounded models assessing the ability of the SNPs to differentiate individuals based on their genetic similarity to global superpopulations performed better than models predicting OUD status (Figure 3 and Supplementary Table 9). Models distinguishing individuals who were genetically similar to EUR from AFR superpopulations had an accuracy of 90.22% (95% CI = 89.63 to 90.79%) and those distinguishing AFR from AMR had an accuracy of 87.53% [95% CI = 86.56 to 88.46%]. The model was less accurate at distinguishing EUR from AMR (66.07% [95% CI = 65.15 to 66.99%]), likely due to the higher prevalence of genetic similarity between EUR and AMR individuals.^25^

**Figure 3.**
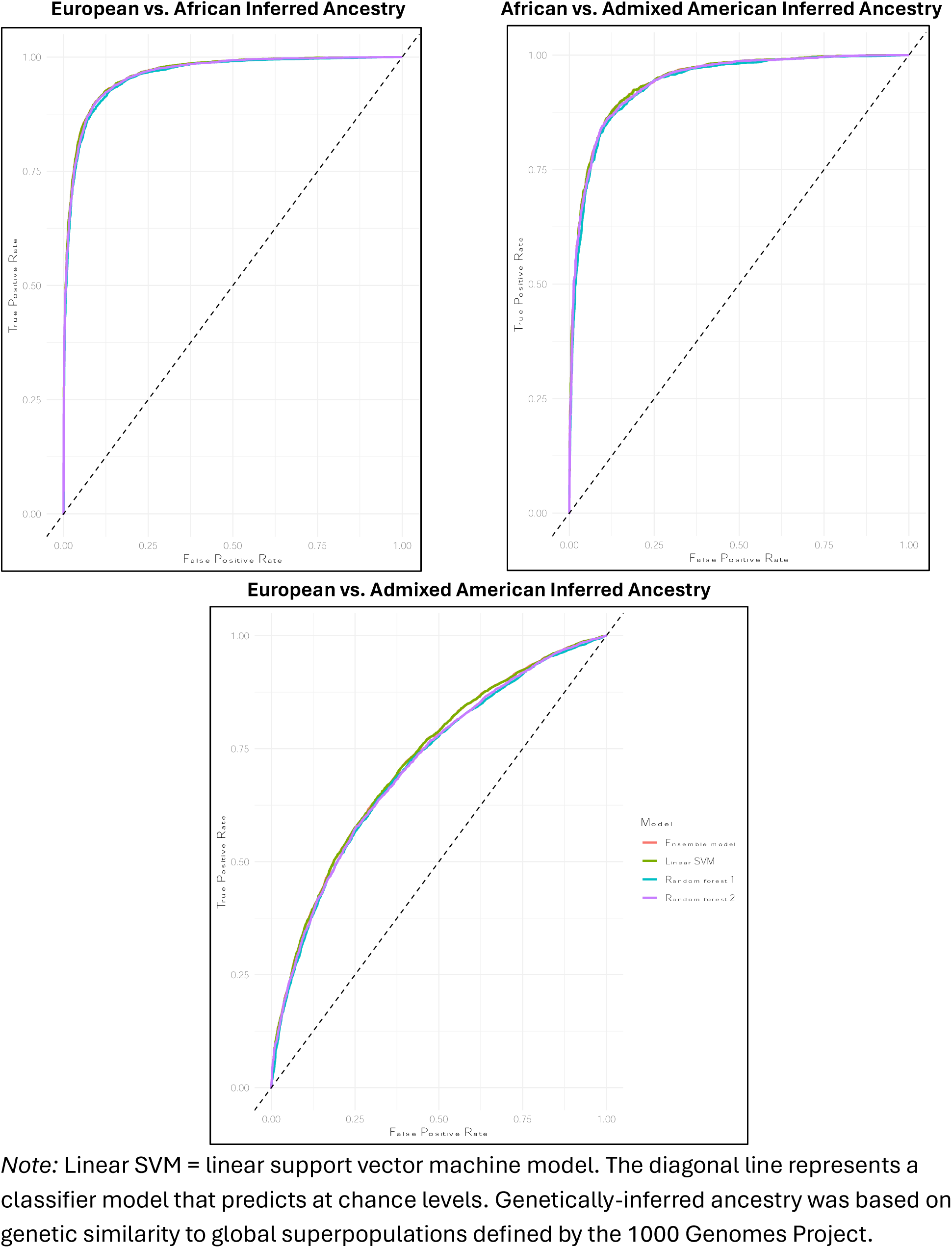
Receiver operating characteristic (ROC) curves of models predicting genetically-inferred ancestry from 15 candidate single nucleotide polymorphisms.

In the full sample ML models, age and sex alone yielded more accurate predictions of OUD risk (59.49% [95% CI = 58.82 to 60.16%]) than the 15 SNPs. The sensitivity and specificity of the model were 0.51 and 0.68, respectively. The accuracy of models within GIA groups was comparable for individuals who were genetically similar to AFR and AMR superpopulations (AFR: 57.92% [95% CI = 56.50 to 59.33%]; AMR: 57.78% [95% CI = 55.32 to 60.21%]), and better in individuals who were genetically similar to the EUR superpopulation (63.50% [95% CI = 62.55 to 64.43%]). Full results are in Supplementary Tables 10 and 11.

## Discussion

In a diverse sample of over 450,000 opioid-exposed individuals (including 33,669 OUD cases), we found no evidence to support the clinical utility of the 15 candidate SNPs purported to predict OUD risk using a proprietary algorithm. Collectively, the SNPs accounted for 0.4% of the variance in OUD risk, consistent with small individual effects of common genetic variants on complex traits.^26,27^ In a hold-out testing subsample, we observed high rates of false positives and negatives, with approximately 47 of 100 predicted cases or controls being incorrectly classified, which is no better than random chance prediction. False positives can contribute to stigma, cause patients undue concern, and bias healthcare decisions.^28^ For example, providers exposed to information that emphasized genetic causes of mental health conditions had less empathy toward patients than providers presented with psychosocial causes.^29^ False negatives could give patients and prescribers a false sense of security regarding opioid use and lead to inadequate treatment plans. Notably, clinicians could better predict OUD risk using an individual’s age and sex than the 15 genetic variants. In summary, although the test approved by the FDA is intended to complement standard clinical assessment, its use is unlikely to confer additional benefits and may instead give providers and patients false and potentially harmful information.

We also found consistent evidence that when variations in genetic similarity were not considered, it resulted in biased findings. In single-SNP logistic regressions, 10 of 13 significant associations (77%) with OUD risk were rendered nonsignificant when measures of global genetic similarity were included as covariates. Half of these associations showed a reversal in the direction of effect when genetic similarity covariates were included. In contrast, the ML models highlighted the SNPs’ ability to differentiate individuals who varied in their genetic similarity to global superpopulations with high accuracy, implying ancestral confounding in unadjusted models.^14^ To avoid spurious associations and ensure the validity of results, developers of genetic risk models intended for clinical care should carefully consider and account for bias attributable to population stratification and sociodemographic factors. Failure to do so may exacerbate existing health inequities.^30^

Several limitations of this study should be considered. First, models were evaluated using EHR diagnosis codes, whose assignment is susceptible to bias.^31^ However, comparable results were previously obtained in a smaller sample where diagnoses were assigned using structured interviews.^14^ Second, the MVP sample is predominantly male, though given its size, the analyses included over 40,000 women (2,619 OUD cases), which far exceeds the full sample on which the pre-marketing approved algorithm was developed (N_total_ = 1,762, N_cases_ = 653; EM-24069 AvertD™ Package Insert as of 12-15-2023). The MVP sample also has higher rates of OUD and pain^32^ and is older than the general population.^33^ We encourage efforts to evaluate the 15 genetic variants in additional datasets. Third, we used GIA group–the current standard for MVP research–as a population descriptor. Despite being an improvement over previous descriptors that combined GIA with self-reported race and ethnicity,^34^ we note that GIA population descriptors do not fully align with recent guidance.^21^ Finally, MVP uses array genotyping, which is less accurate than, for example, that involving mass spectrometry,^35^ and imputation was required for about half of the SNPs. Although this may have reduced genotyping accuracy, which is important for individual-level risk prediction, it would not be expected to impact the results in a large sample like ours.

Genetics researchers have argued against the use of candidate genes to predict OUD and other psychiatric traits.^36^ Because genetic risk models in psychiatry will continue to emerge and could prove to be clinically useful, it is crucial that researchers and regulatory agencies adopt rigorous standards for developing and evaluating them prior to their application in clinical settings. When regulatory agencies evaluate genetic risk algorithms that use advanced statistical methodologies (e.g., ML), it is imperative to heed the guidance of scientific advisors and independently validate the findings. By applying rigorous standards to reduce sources of bias, the potential benefits of genetic risk models can be maximized while protecting patient safety and well-being.

## Supporting information

Supplementary Acknowledgments

## Data Availability

Individual-level Million Veteran Program data is available to approved Veterans Affairs researchers.

## Acknowledgments

This research is based on data from Million Veteran Program, Office of Research and Development, Veterans Health Administration and was supported by awards #I01 BX003341 and IK2 CX002336 (to EEH); the VISN 4 Mental Illness Research, Education, and Clinical Center; NIAAA grant K01 AA028292 (to RLK); and NIDA grant P30 DA046345. An MVP Core Acknowledgement list is provided in the supplemental materials. This publication does not represent the views of the Department of Veteran Affairs, the Department of Defense, Uniformed Services University, or the United States Government.

## Notes

### Competing Interest Statement

Dr. Kranzler is a member of advisory boards for Dicerna Pharmaceuticals, Sophrosyne Pharmaceuticals, Enthion Pharmaceuticals, and Clearmind Medicine; a consultant to Sobrera Pharmaceuticals; the recipient of research funding and medication supplies for an investigator-initiated study from Alkermes; and a member of the American Society of Clinical Psychopharmacology Alcohol Clinical Trials Initiative, which was supported in the last three years by Alkermes, Dicerna, Ethypharm, Lundbeck, Mitsubishi, Otsuka, and Pear Therapeutics, Drs. Kranzler and Gelernter hold U.S. patent 10,900,082 titled: "Genotype-guided dosing of opioid agonists," issued 26 January 2021.

### Author Declarations

The Million Veteran Program (MVP), an initiative of the U.S. Department of Veterans Affairs to discover the genetic etiology of diseases relevant to Veterans, was approved by the Central Veterans Affairs Institutional Review Board (IRB) and all site-specific IRBs. We followed all relevant ethical regulations for research with human subjects.

