## Supplementary Figure 2 for "Candidate Genes from an FDA-Approved Algorithm Fail to Predict Opioid Use Disorder Risk in Over 450,000 Veterans"

**Supplementary Figure 2.** Scatter plot of the first two ancestry principal components among opioid exposed individuals.

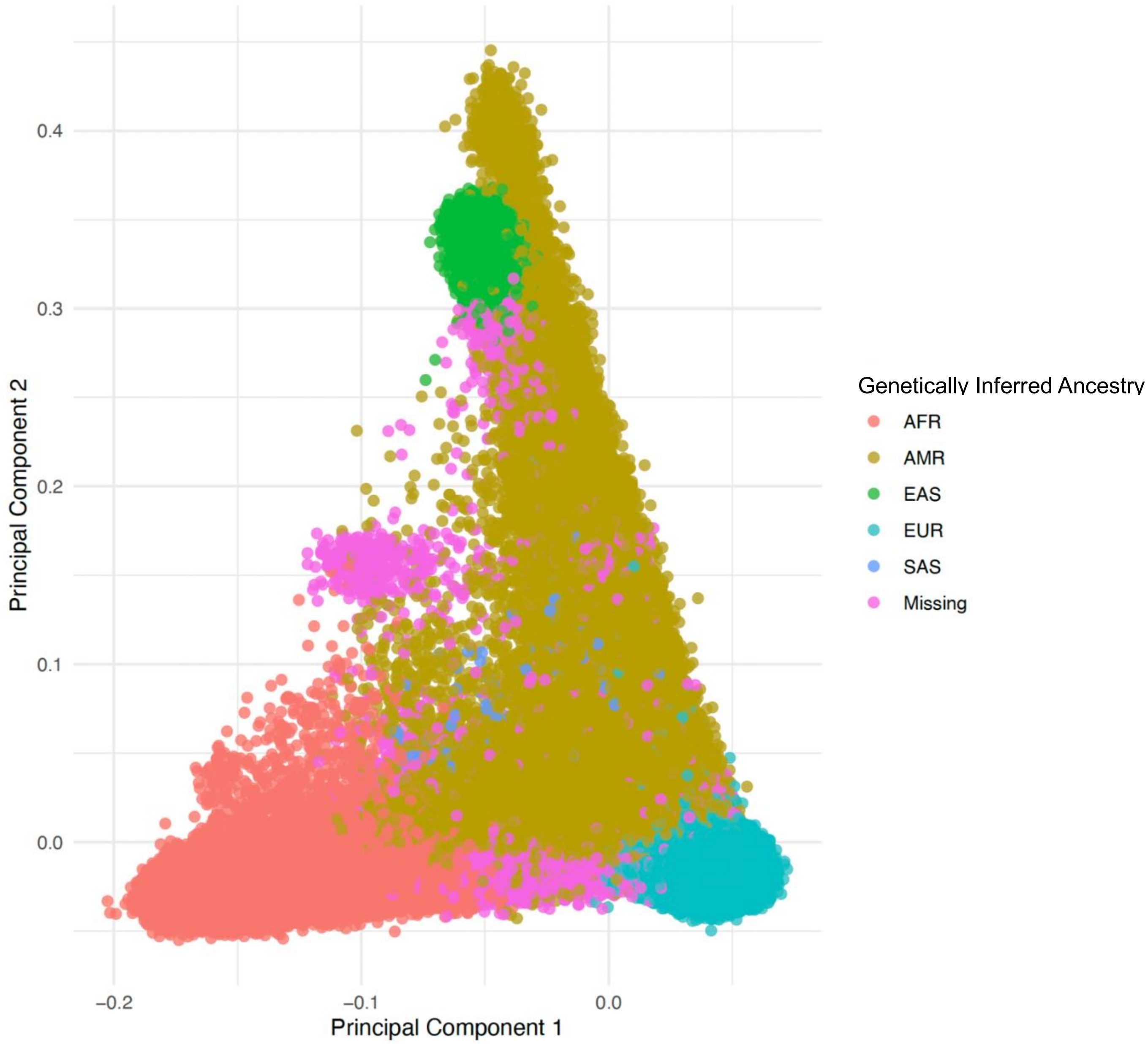


AFR = individuals genetically similar to the African superpopulation, AMR = individuals genetically similar to the admixed American superpopulation, EAS = individuals genetically similar to the East Asian superpopulation, EUR = individuals genetically similar to the European superpopulation, SAS = individuals genetically similar to the South Asian superpopulation. Individuals missing on genetically inferred ancestry were unable to be classified with a predicted probability over 50% to any given cluster based on their genetic similarity.
