## Supplementary Figure 1 for "Candidate Genes from an FDA-Approved Algorithm Fail to Predict Opioid Use Disorder Risk in Over 450,000 Veterans"

**Supplementary Figure 1.** Alternate allele frequency across inferred ancestry groups in the Million Veteran Program participants.

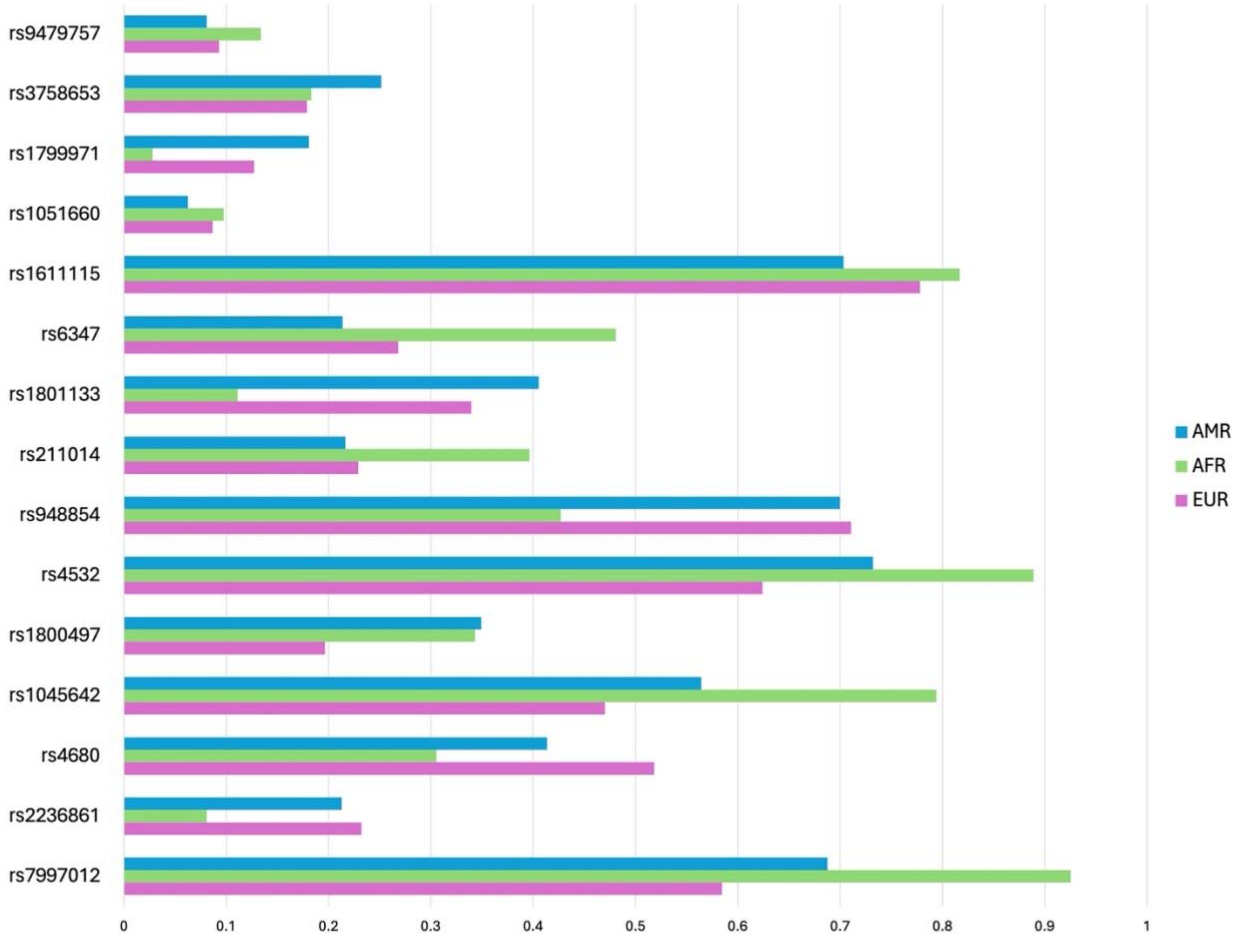


Inferred ancestry was assigned based on genetic similarity to superpopulations defined by the 1000 Genomes Project. AMR = admixed American, AFR = African, EUR = European.
